# Comparing traditional, computerised and virtual reality assessments of social cognition in schizophrenia: A within-subjects multimodal approach

**DOI:** 10.1101/2025.05.09.25327050

**Authors:** Kirsten Gainsford, Bernadette M. Fitzgibbon, Aron T. Hill, Paul B. Fitzgerald, Caroline T. Gurvich, Kate E. Hoy

## Abstract

Social cognition is significantly impacted in people with schizophrenia and can be assessed using various methods including traditional (paper-and-pencil) tasks, computer tasks, and Virtual Reality (VR) assessments. The current study investigated whether these different approaches to social cognitive assessment, with a particular focus on theory of mind (ToM), could consistently and sensitively identify differences between individuals with schizophrenia and controls within the same sample. We hypothesised that participants with schizophrenia would perform less well than controls across all assessment methods. We additionally measured brain changes associated with social cognition during the ToM computer task using electroencephalography (EEG). Our results revealed that the schizophrenia group performed less well than the control group in ToM across all assessment approaches. They also performed less well on traditional measures of social knowledge and on VR measures emotion recognition. Additionally, event-related potential (ERP) amplitudes were reduced in people with schizophrenia compared to controls across brain regions linked to ToM. Our findings suggest that these different modes of social cognitive assessment are all sensitive to detecting differences in ToM abilities, with VR in particular showing strongest effect sizes. Implications of the findings on future protocol development are discussed.

## 1. Introduction

Social cognition broadly refers to the ability to understand, interact with and interpret the social environment and the people around us (Green et al., 2008). Social cognitive skills are separated into several inter-related domains, including theory of mind (ToM; the ability to infer what others’ intentions and beliefs are and to make meaning of them), emotion recognition (identifying, perceiving and understanding emotions), social knowledge (awareness of societal rules, etiquette and roles in social interaction), and attributional bias (how people assign cause to positive and negative events that occur, whether it be to the self, other or another external factor; Green et al., 2008). Social cognition has an impact on functioning across the lifespan (Tandon et al., 2024) and poor social cognitive skill development can have a significant impact on daily functioning (Green, 2016). There are well-documented difficulties with social cognition in people with schizophrenia (Green, 2016), which has been associated with reduced ability to obtain and maintain jobs (Holm, Taipale, Tanskanen, Tiihonen, & Mitterdorfer-Rutz, 2021), to develop close relationships with others (Green, 2016), to manage conflicts (Buck, Browne, Gagen, & Penn, 2023), and overall reduced functioning and quality of life (Tandon et al., 2024; Thibaudeau, Cellard, Turcotte, & Achim, 2021).

Over time, a number of tools have been developed to measure these different social cognitive domains (Eddy, 2019; Pinkham, Harvey, & Penn, 2018; Tsui et al., 2024), traditionally via paper-and-pencil or computer-based tasks. Traditional measures require participants to make social judgements and inferences from static, non-naturalistic stimuli (Bohil, Alicea, & Biocca, 2011; Teufel et al., 2013) and are often carried out in sterile clinical or research environments (Horan et al., 2011; Parsons, 2015; Parsons, Gaggioli, & Riva, 2017). When comparing the psychometric properties of these tasks, the criteria researchers use often differ and thus the conclusions about assessment quality can vary significantly (Pinkham et al., 2018; Tsui et al., 2024). While some researchers may focus on measuring task quality via consistency and sensitivity, others focus on generalisability or accuracy. For example, a recent large-scale review of social cognitive tasks concluded that ToM assessment the Hinting Task was not a satisfactorily consistent measure (Tsui et al., 2024). However, another review identified the Hinting Task as the ToM task with the strongest psychometric properties for use in clinical trials (Pinkham et al., 2018). Traditional measures also have the potential of administrator confounds or biases such as with scoring and standardisation of instructions (Hoyt, 2000; Uher, 2022).

Computer-based tasks were introduced to improve on traditional tasks and have shown comparable results with them (Chan, Kwong, Wong, Kwok, & Tsoi, 2018; Friedman, Kohler, Gunalp, Boone, & Hegarty, 2020). They remove administrator error by automatically managing randomisation of stimuli presentation, accurately enforcing test timing limits, and calculating scores quickly and simply (Friedman et al., 2020), thus improving rigorous control of the measures and thereby, improving consistency, reliability and sensitivity (Bailey, Neigel, Dhanani, & Sims, 2018; Chan et al., 2018). Computer based tasks are also conducive to measurement of brain activity associated with the skills being tested (i.e., social cognition) via technologies such as electroencephalography (EEG). Combining EEG with social cognitive measures can help characterise brain responses such as event-related potentials (ERPs) providing insights into the underlying neuropathophysiology of disorders with reduced social cognitive skills such as schizophrenia (Li et al., 2025). In schizophrenia-related research investigating ERP differences in schizotypy on ToM tasks, a schizotypy group had reduced ERP amplitudes in response to emotional ToM stimuli at the right temporoparietal junction (rTPJ), a ToM processing hub (Leung, Lei, Wang, & Lam, 2021). In broader ERP ToM research, the TP450, peaking at 450ms after stimuli presentation and in temporoparietal regions such as the rTPJ, has been shown to be activated in ToM tasks requiring taking others’ perspectives. It has also been suggested as a marker of the time course of ToM processing (Gan et al., 2016; Peng et al., 2018; Vistoli et al., 2015). These findings support for the use of neurophysiological techniques alongside behavioural measures to measure ToM related brain activity.

While EEG can provide additional insights in conjunction with computer tasks, a significant criticism of computer and traditional tasks is that they can oversimplify the measurement of some complex thought processes and skills, as well as still taking place in environments which lack ecological validity. All of which impacts the transfer of results to real-world applications (Parsons et al., 2017). Virtual reality (VR) is an easily accessible, relatively affordable and immersive tool with high ecological validity (Parsons, 2015; Parsons et al., 2017) that is being more frequently utilised for assessment and treatment in schizophrenia for this reason (Hoşgelen, Güneri, Erdeniz, & Alptekin, 2024). VR is particularly well suited to the assessment of social cognition as participants can be placed in virtual environments where social interactions are usually encountered such as at shopping malls, restaurants and bus stops (Canty, Neumann, Fleming, & Shum, 2015) and evaluate how they conduct themselves in these more realistic social scenarios. VR can also be used to re-create traditional cognitive and social cognitive assessments, such as the “so-moral” task used to measure sociomoral decision making, in a less clinical environment (Morasse, Vera-Estay, & Beauchamp, 2021). Similarly to the traditional methods, these VR tasks have shown sensitivity in being able to successfully differentiate between schizophrenia and control groups (Rus-Calafell, Garety, Sason, Craig, & Valmaggia, 2018).

As demonstrated, there are a multitude of ways in which to assess these complex social cognitive skills, including ToM. Often, comparisons of assessment approaches are completed between samples and across studies (Tsui et al., 2024). It may be useful, instead, to measure consistency across assessments to compare them within the same sample. Doing this could provide more certainty that observed differences in ToM skills are robust findings and as a result, it may also ensure confidence that when using a particular assessment approach, it is suited to the context, the patient and the researcher.

The current study aimed to investigate whether a multi-method assessment approach of traditional, computer-based, and VR tasks would consistently detect differences between a schizophrenia and control group on ToM skills, within the same sample. We also compared changes in ERP amplitude between schizophrenia and control groups during ToM computer task performance to differentiate brain activity responses to ToM stimuli at ToM-relevant brain areas (i.e., rTPJ). We hypothesised that the schizophrenia group would perform more poorly than controls on ToM tasks across all assessment approaches. We also hypothesised that the schizophrenia sample would have a lower TP450 ERP peak amplitude during a ToM computer task at the rTPJ compared to controls.

## 2. Methods

### 2.1 Participants

A total of 21 healthy controls and 17 people with a diagnosis of schizophrenia (n = 10) or schizoaffective disorder (n = 7), were enrolled into the study. Two participants with a diagnosis of schizophrenia or schizoaffective disorder withdrew: one after consent, one after partial completion of data collection. Participants with incomplete data were excluded from analysis. Therefore, the final analysed sample consisted of 36 individuals (15 [9F, 6M] schizophrenia [9 schizophrenia, 6 schizoaffective disorder]; 21 controls [12F, 9M]; see **Table 1** for demographics).

**Table 1:**
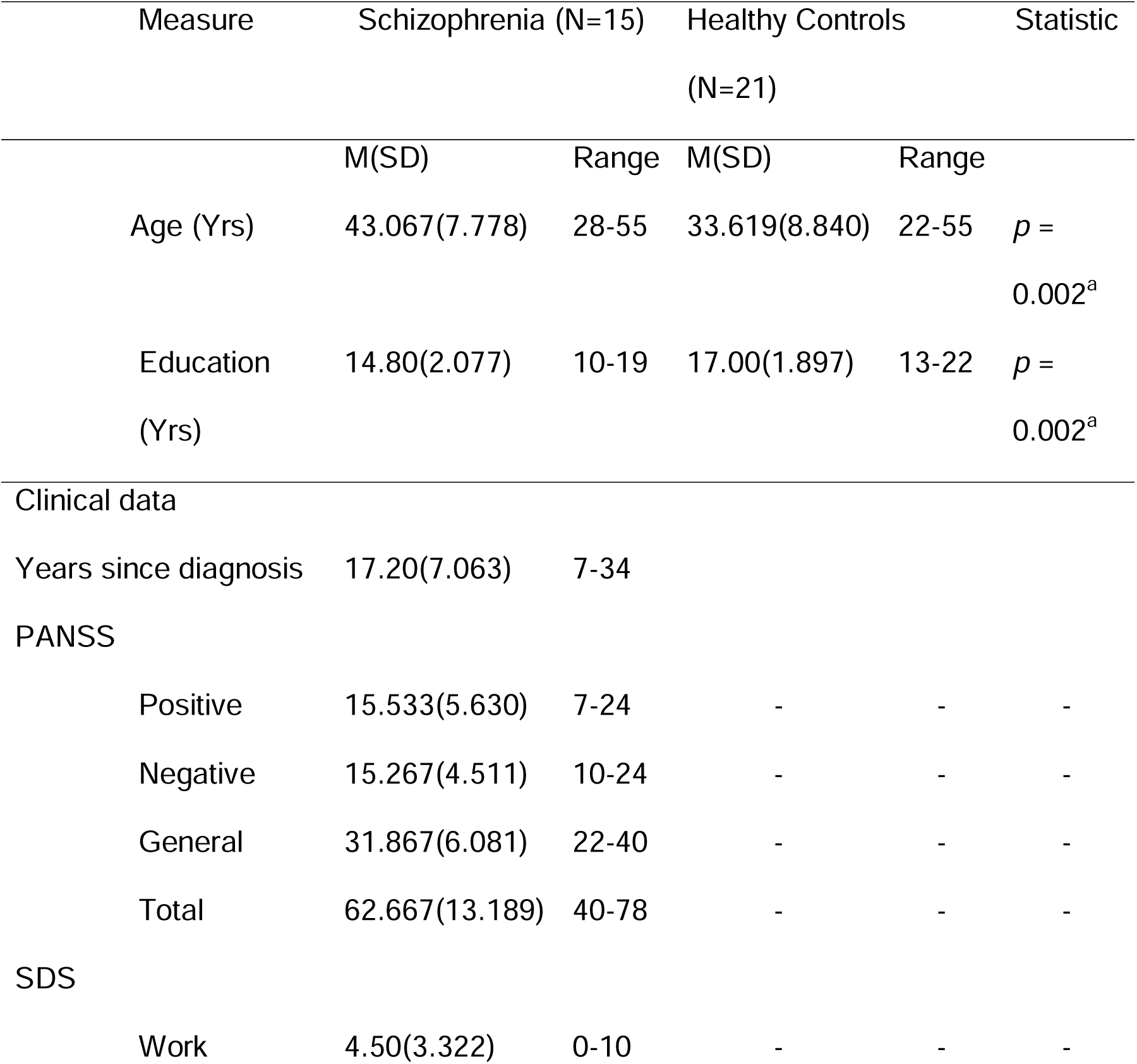
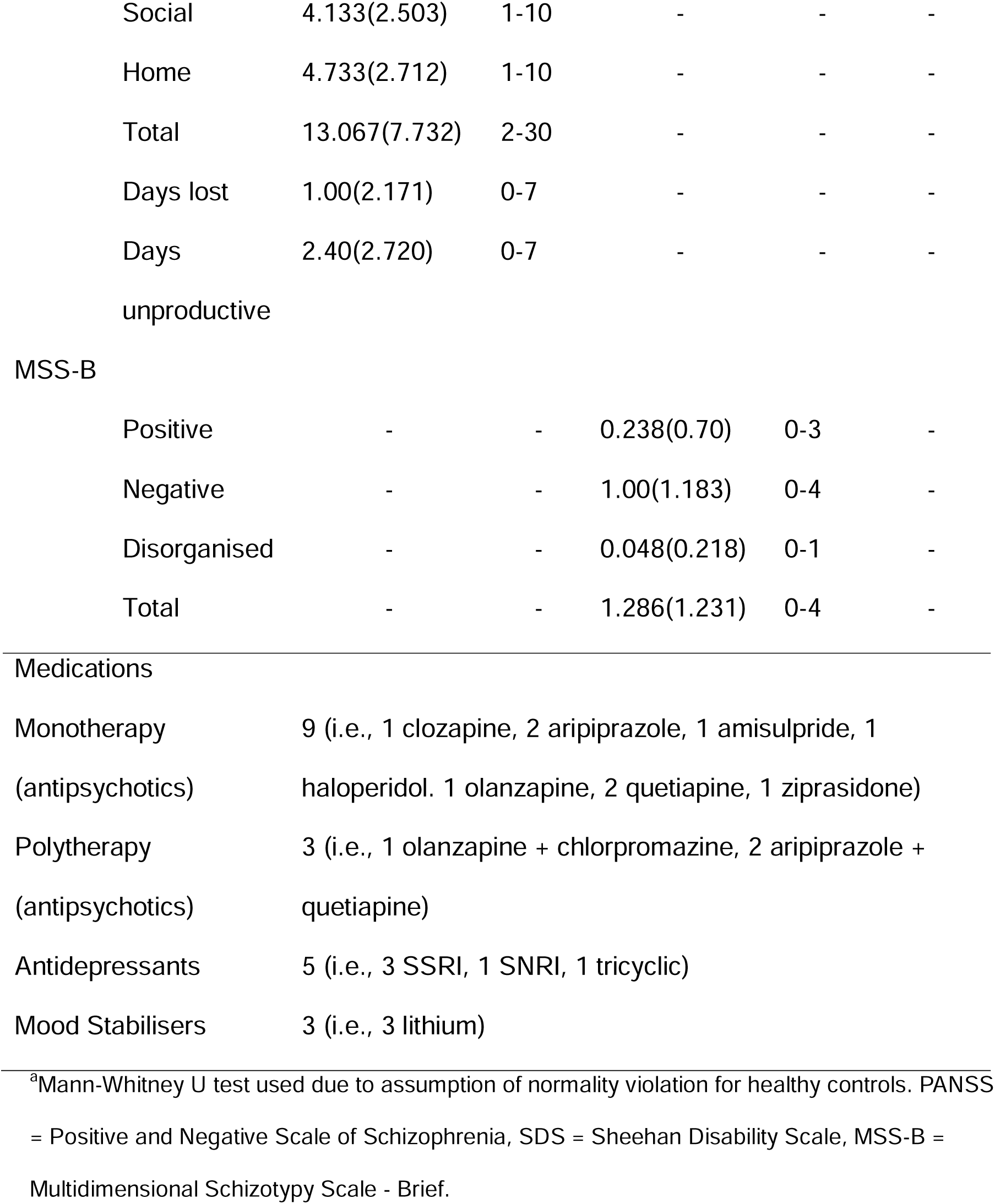
Participant Descriptives.

| Measure | Schizophrenia (N=15) |  | Healthy Controls<br>(N=21) |  | Statistic |
| --- | --- | --- | --- | --- | --- |
|  | M(SD) | Range | M(SD) | Range |  |
| Age (Yrs) | 43.067(7.778) | 28-55 | 33.619(8.840) | 22-55 | $p = 0.002^a$ |
| Education<br>(Yrs) | 14.80(2.077) | 10-19 | 17.00(1.897) | 13-22 | $p = 0.002^a$ |
| Clinical data |  |  |  |  |  |
| Years since diagnosis | 17.20(7.063) | 7-34 |  |  |  |
| PANSS |  |  |  |  |  |
| Positive | 15.533(5.630) | 7-24 | - | - | - |
| Negative | 15.267(4.511) | 10-24 | - | - | - |
| General | 31.867(6.081) | 22-40 | - | - | - |
| Total | 62.667(13.189) | 40-78 | - | - | - |
| SDS |  |  |  |  |  |
| Work | 4.50(3.322) | 0-10 | - | - | - |
| Social | 4.133(2.503) | 1-10 | - | - | - |
| Home | 4.733(2.712) | 1-10 | - | - | - |
| Total | 13.067(7.732) | 2-30 | - | - | - |
| Days lost | 1.00(2.171) | 0-7 | - | - | - |
| Days unproductive | 2.40(2.720) | 0-7 | - | - | - |
| MSS-B |  |  |  |  |  |
| Positive | - | - | 0.238(0.70) | 0-3 | - |
| Negative | - | - | 1.00(1.183) | 0-4 | - |
| Disorganised | - | - | 0.048(0.218) | 0-1 | - |
| Total | - | - | 1.286(1.231) | 0-4 | - |

Medications
|  |  |
| --- | --- |
| Monotherapy<br>(antipsychotics) | 9 (i.e., 1 clozapine, 2 aripiprazole, 1 amisulpride, 1 haloperidol, 1 olanzapine, 2 quetiapine, 1 ziprasidone) |
| Polytherapy<br>(antipsychotics) | 3 (i.e., 1 olanzapine + chlorpromazine, 2 aripiprazole + quetiapine) |
| Antidepressants | 5 (i.e., 3 SSRI, 1 SNRI, 1 tricyclic) |
| Mood Stabilisers | 3 (i.e., 3 lithium) |

<sup>a</sup>Mann-Whitney U test used due to assumption of normality violation for healthy controls. PANSS = Positive and Negative Scale of Schizophrenia, SDS = Sheehan Disability Scale, MSS-B = Multidimensional Schizotypy Scale - Brief.

Schizophrenia and schizoaffective disorder diagnosis was confirmed using the Mini-International Neuropsychiatric Interview (MINI; Sheehan, 2016) administered by a trained researcher (KG). Participants with schizophrenia were all living in the community at the time of participation. Twelve of these participants were regularly taking one or more antipsychotic medications (see **Table 1**), one person was receiving only psychotherapeutic treatment, two people were not receiving treatment. All participants taking medications were on stable doses in the four weeks prior to enrolling in the study and for the duration of their participation in the study.

The data reported in the current paper was collected as part of a larger study investigating the effects of brain stimulation on social cognition which necessitated the following criteria be met for participation: for healthy controls, the exclusion criteria were any history of diagnoses of mental health conditions. Exclusion criteria for both groups included any unstable medical or neurological conditions, any substance use disorders or other significant psychiatric comorbidities as well as any significant traumatic brain injury, history of seizure or epilepsy. Participants were also excluded if they were pregnant or breastfeeding.

Data collection took place at two sites, Epworth Centre for Innovation in Mental Health (ECIMH) during 2021-2022 and Monash Alfred Psychiatry Research Centre (MAPrc) during 2022-2023. All equipment used was the same at both sites and all participants completed their full set of data collection at a single site.

Participants were recruited via recruitment websites (e.g., HealthMatch), social media (e.g., Facebook, X), referrals from the team’s participant database (consisting of participants that had previously participated in our research that consented to be contacted for participation opportunities in other studies) as well as referrals from community support services (e.g., hospitals, GP and psychiatry clinics, social support services and housing care). Participants were also recruited via flyers and at community centres. All participants provided written informed consent. Ethics approval was obtained from the Monash Health Human Research Ethics Committee and governance obtained through Monash University, Alfred Health and Epworth HealthCare.

### 2.2 Design

As described above, data was collected as part of a larger study investigating the effects of brain stimulation on social cognition, however, only baseline (pre-stimulation) data are reported here. Baseline assessments included a battery of traditional cognitive and social cognitive tasks as well as a computer task and VR social cognitive tasks. The schizophrenia group completed a clinical measure (Positive and Negative Schizophrenia Scale - PANSS; Kay, Opler, & Lindenmayer, 1989) and controls completed the Multidimensional Schizotypy Scale (MSSB-B; Gross, Kwapil, Raulin, Silvia, & Barrantes-Vidal, 2018). Participants also underwent EEG during the completion of a ToM intention attribution computer task. Participants attended two testing sessions each.

### 2.3 Measures

All assessments were conducted by a trained researcher (KG).

#### 2.3.1 Cognitive Measures

Standardised cognitive tasks included sub-tests from the MATRICS Consensus Cognitive Battery (MCCB; Nuechterlein et al., 2008). These included, Trial making task A, Brief Assessment of Cognition in Schizophrenia: Symbol Coding (BACS-SC), Brief Visuospatial Memory Test-Revised (BVMT-R), Category Fluency: Animal Naming and the Hopkins Verbal Learning Test-Revised (HVLT-R). Additionally, the Digit Span forward and backwards tests from the Wechsler Adult Intelligence Scale (WAIS-IV; Drozdick, Wahlstrom, Zhu, & Weiss, 2012) were used. These tasks were administered according to standardised instructions.

#### 2.3.2 Theory of Mind Measures

Theory of Mind was assessed across all three modalities, namely traditional, computer based and VR.

##### 2.3.2.1 Traditional (i.e., paper and pencil) Task

The Abbreviated Faux Pas test was extracted from the Mini Social Cognitions and Emotion Assessment (Mini-SEA; Bertoux et al., 2012) to test ToM. This version of the task has 10 comic strips created based on the original Faux Pas scripts. There are five faux pas and five non-faux pas comics. Participants read the comics and answer the same series of questions required in the original Faux Pas test (Gregory et al., 2002; Stone, Baron-Cohen, & Knight, 1998). A sub-score out of 15 was calculated based on Mini-SEA instructions and used for analysis (Bertoux et al., 2012).

##### 2.3.2.2 Computer-Based Task

Participants completed the attribution of intentions task used in work by Vistoli et al. (2015). The task consists of a series of comic strips, each containing four images. There are three types of comic strips each with a congruent and incongruent ending option. The first is the attribution of intentions comics which require the use of ToM to interpret. The second and third comic types, act as control stimuli. The second type of comic includes human characters that cause physical rules of the environment such as gravity to take place (physical causality with character – PCCh). The third type of comic included only objects and physical causality (PCOb). The two latter comic types do not require ToM to interpret. In this study, the AI comics were presented separately from the PCCh and PCOb comics. The latter two sets of comics were combined to form a non-theory of mind (NToM) comic condition. The ToM task contained 24 comics and the NToM task contained 48 comics. There were two versions of each type of comic (i.e., ToM A, ToM B, NToM A, NToM B). Whether comics had a congruent or incongruent ending was randomised and comic order within each task was randomised task version was counterbalanced, and participants saw each version of the task once in the session.

To complete the tasks, participants sat in a chair approximately one metre from the computer screen. EEG recording was conducted simultaneously while participants completed the task (see **Section 2.4** for EEG details). For the ToM task, participants pressed a green button on a button box if the fourth picture in the comic “makes sense for the person’s goals” and a red button if it did not. For the NToM task, participants pressed the green button if the fourth picture “makes sense as the ending of the story” and a red button if it did not. **Figure 1** shows the time sequence of the comics. The fourth picture was presented until the participant responded or for 4000ms, whichever came first. To assist participants in knowing when to respond, the fourth picture of each comic had a red square around it (**Figure 1**). Number of correct responses and response times were recorded.

**Figure 1:**
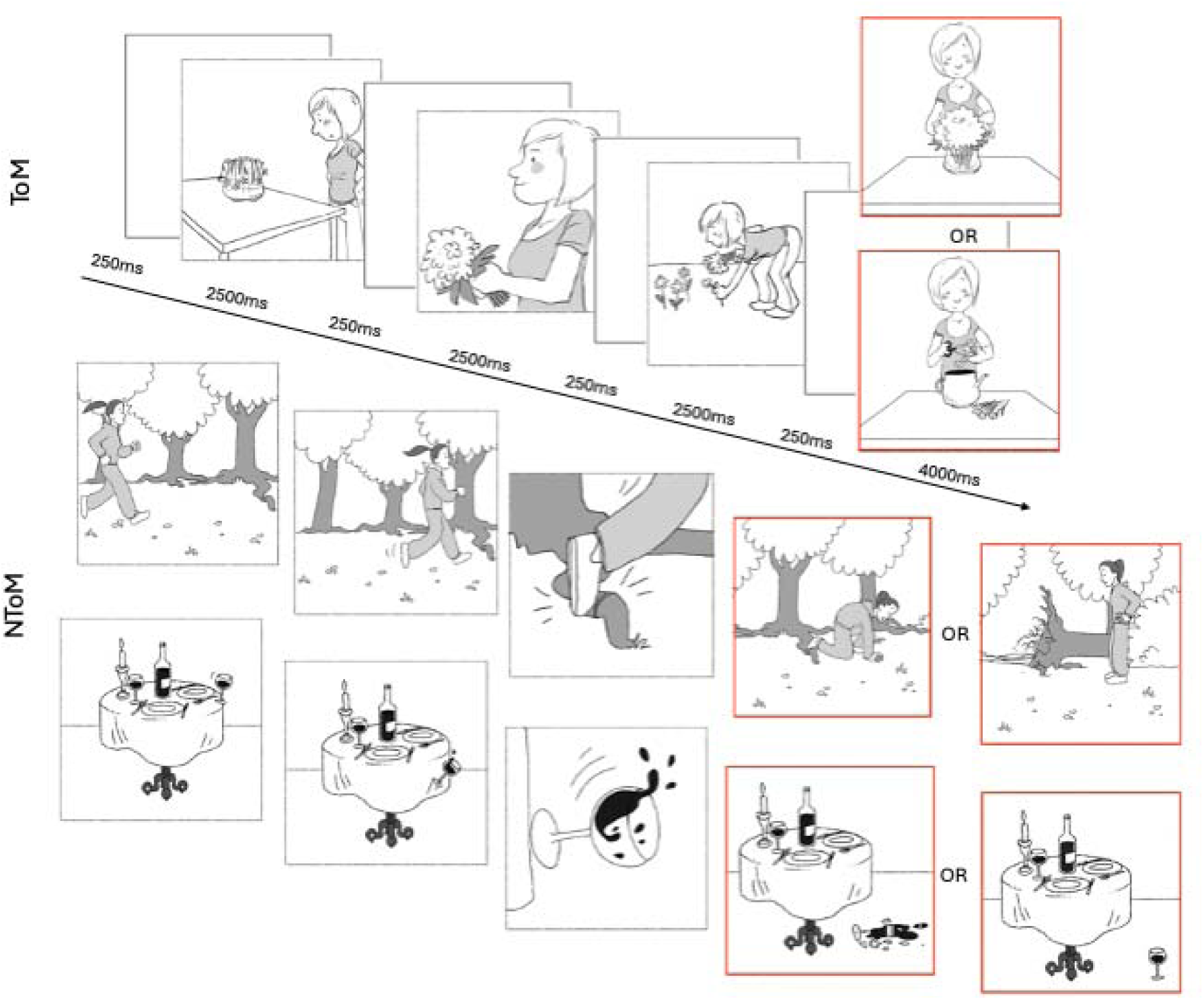
Attribution of intentions ToM task. The first is a ToM example with congruent or incongruent ending options and NToM examples (PCCh and PCOb) with alternate endings below.

##### 2.3.2.3 Virtual Reality Tasks

To measure social cognition in a virtual environment, participants wore a Meta Quest 2 head-mounted display with hand-held controllers (Meta, 2020). They were placed in a virtual living room with comfortable, homely features. Participants sat on a virtual couch and completed the tasks on the television in the virtual living room. A virtual avatar explained the instructions of each task to the participant before starting. To complete the task, the participant pointed the right-hand controller at the television, highlighted the answer they would like to select, and pulled the trigger (SoCog, SensiLab, Monash University, Melbourne, Australia).

To measure ToM, the Reading the Mind in the Eyes Task (RMET, Baron-Cohen, Wheelwright, Hill, Raste, & Plumb, 2001) and Hinting task (Corcoran, Mercer, & Frith, 1995) were adapted to be viewed on the virtual television, the same way it would be presented on a computer screen (**Figure 2**). The RMET used the same 36 stimuli and showed a set of eyes on the screen and head to determine what the person was thinking or feeling. There were four options to select from underneath the eyes. If participants required definitions of the options provided, the researcher explained them. Participants completed all 36 trials and then if any were incorrect the first time around, they were repeated. The program was set up so that this continued until all responses were correct. However, for the purpose of this research, only the first response to each trial was used in analysis.

**Figure 2:**
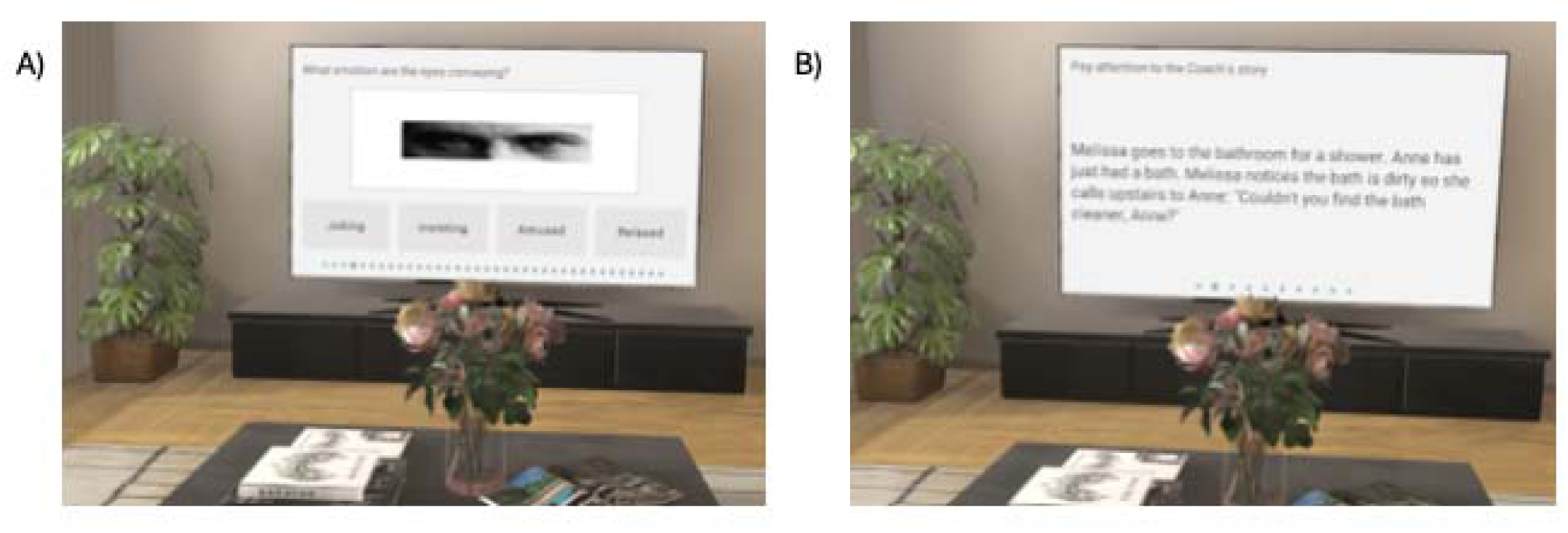
Example of VR ToM tasks completed in virtual living room on the television. Participants completed the “Reading the Mind in the Eyes Task” (RMET, Figure 2A) and Hinting Task (Figure 2B) using the right-hand controller as a laser pointer and pulled a “trigger” on the controller to respond.

The 10 Hinting Task vignettes were read out to the participant by the virtual avatar, and they could select if they wanted the story repeated to them. Participants first completed all 10 questions and then any incorrect ones were repeated. Only the first attempt at each of the 10 questions was used in analysis.

#### 2.3.3 Emotion Recognition, Social Knowledge and Attributional Bias Tasks

Emotion recognition was measured using VR. Social knowledge and attributional bias were measured using traditional assessments.

Faces from the NimStim facial expressions database (Tottenham et al., 2009) were used to build a four-stage VR emotion recognition task. The images used were male and female from diverse cultural backgrounds. The images showed happy, angry, sad, fearful, surprised, disgust, calm and neutral expressions. The specific instructions for each stage of the task were given by the virtual avatar. Participants first completed each question of the stage and then any incorrect questions were re-done until they were all correct. If participants did not get enough questions correct the first time, the program required them to re-do that stage of the task. If this was required, the researcher provided the responses for that stage to allow the participant to move to the next stage. Only the first response to each question in each stage was required and therefore used for analysis. Stage 1 had 15 trials and showed a single image of a face and asked, “Please select the correct emotion” and provided four text options to select from. Stage 2 had 16 trials and showed four images of facial expressions of the same person and asked, “which face shows [insert emotion]?”. Stage 3 had 16 trials and showed two images side by side and required participants to respond as quickly as they could to the question “which face shows [insert emotion]?” by selecting the face that showed the mentioned. Stage 4 had 16 trials and showed a blurred image of a face and asked, “please select the correct emotion” and provided four text options to select from. Participants completed the task in the same virtual living room environment described in **Section 2.3.2.3**.

Traditional tasks were used to measure social knowledge and attributional bias. The Internal, Personal and Situational Attributions Questionnaire (IPSAQ) was used to measure attributional bias (Kinderman & Bentall, 1996) and social knowledge was measured using the Situational Feature Recognition Test (SFRT; Corrigan & Green, 1993). The two latter assessments were also administered per standardised task instructions.

### 2.4 EEG

EEG was recorded via Brain Vision Recorder software (Brain Products, Gilching, Germany) with a 50-electrode montage using the 10-05 system (ActiCap Slim, Brain Products, GmbH, Gilching, Germany). Electrodes (Ag/AgCl) were connected to a BrainAmp amplifier (Brain Products, GmbH, Gilching, Germany). The electrodes used were AF3, AFz (ground), AF4, F7, F5, F3, F1, Fz, F2, F4, F6, F8, FC5, FC3, FC1, FCz, FC2, FC4, FC6, T7, C5, FCC6h, CCP5h, C3, C1, Cz, C2, C4, FCC6h, CCP6h, T8, TP7, CP5, CP3, CP1, CPz (reference), CP2, CP4, CP6, TP8, P7, P5, P3, P1, Pz, P6, P8, PO3, POz, PO4, O1, Oz and O2. The electrode montage was customised to accommodate for transcranial alternating current stimulation (tACS) electrodes as well as use of VR with the cap on (for use as part of the larger study). Impedances were regularly checked and kept below 10 kΩ throughout the EEG session. Data were collected at a sampling rate of 1000Hz with a high-pass filter of 0.01Hz and low-pass filter of 200Hz.

### 2.5 Statistical Analysis

#### 2.5.1 Cognitive Measures

A series of independent sample t-tests were conducted to compare cognitive scores between the participants with schizophrenia and the healthy controls using JASP (Version 0.17.3, JASP Team, Amsterdam, Netherlands 2023). In cases where assumptions of normality or equality of variances were violated, non-parametric tests were used.

#### 2.5.2 Social Cognitive Measures

The standardised scores were calculated for each task as specified by scoring instructions for both the VR and traditional measures. However, because this study did not use the full Mini-SEA, only the abbreviated faux pas test sub-score was calculated as per standardised instructions. Additionally, the VR emotion recognition task scoring took an average of the total scores for each of the four task stages and compared the averages between groups. Scores for all assessments were then statistically compared between the schizophrenia and control groups using a series of independent samples t-tests on JASP (Version 0.17.3, JASP Team, Amsterdam, Netherlands 2023). In cases where assumptions of normality or equality of variances were violated, non-parametric tests were used.

The attribution of intentions ToM computer task accuracy was determined by calculating and averaging each participant’s percentage of correct responses. To calculate mean response times, the time at which participants responded to the fourth picture in each comic was subtracted from the total time of each trial and averaged. If participants responded on the button box before the fourth image of the comic was shown (i.e., response error), the trial was excluded from analysis. Independent samples t-tests were then carried out using JASP to compare task accuracy and response time for schizophrenia participants versus control participants.

Pearson’s correlations were also conducted between the traditional, computer-based and VR ToM tasks.

#### 2.5.3 EEG Analysis

All EEG data were pre-processed in MATLAB (2024a, Mathworks, 2024) with the EEGLAB toolbox (Delorme & Makeig, 2004) using RELAX v1.1.5 (Bailey, Biabani, et al., 2023; Bailey, Hill, et al., 2023), a fully automated EEG pre-processing pipeline. The data were first zero-phase Butterworth bandpass filtered. For ERP analysis, the high- and low-pass filters were set at 0.25-80Hz. A 50Hz notch filter was used to remove line noise and the ‘findNoisyChannels’ function from PREP pipeline was used to remove bad electrodes (Bigdely-Shamlo, Mullen, Kothe, Su, & Robbins, 2015). Multi-channel Wiener filtering was then applied to clean muscle activity, horizontal eye movements and blinks and the data were then re-referenced to the common average (Somers, Francart, & Bertrand, 2018). To identify remaining artefacts, independent component analysis (ICA; FastICA-symm method) was performed using the ICLabel classifier (Pion-Tonachini, Kreutz-Delgado, & Makeig, 2019). They were then removed using wavelet enhanced ICA (wICA; Bailey et al., 2024). Finally, rejected electrodes were then re-interpolated using spherical interpolation.

##### 2.5.3.1 Event-Related Potentials

The temporoparietal 450 (TP450) ERP, ranging from 325ms to 525ms and peaking at approximately 450ms in the temporoparietal region, was the focus of analysis. To prepare data for analysis, the above cleaning processes were carried out and data were then epoched. Epochs were created around the third image of each comic as this is the image at which ToM processes are required to understand the storyline in the comic (Vistoli et al., 2015). Epoch lengths for ERP analysis were 1.3 seconds (−300ms to 1000ms). Grand average files of the data were then created using a custom MATLAB (Mathworks, 2024a) script. Region of interest (ROI) analysis was conducted to investigate changes in peak amplitude while participants completed the attribution of intentions task. A group of electrodes representing the right temporoparietal junction (rTPJ) were identified (CP4, CP6, TP8, P6, P8) with the average of the signal across these electrodes used in the analysis. Using JASP, these were statistically compared using a repeated measures ANOVA with mean amplitude during task (ToM and NToM) and group being compared (healthy controls and schizophrenia).

Note, Bayes Factor analyses (BF_10_) were conducted on all non-significant findings.

## 3. Results

### 3.1 Cognitive Assessments

**Table 2** provides a summary of cognitive task data as a characterisation of the samples. A Mann-Whitney U test (due to violation of assumption of normality) showed the schizophrenia group completed the trail making task significantly slower than controls (*U* = 88, *p* = 0.026). The schizophrenia group scored significantly lower on HVLT compared to controls (*U* = 232.50, *p* = 0.016) but not on HVLT delayed recall (BF_10_ = 0.865). The remaining analyses conducted for cognitive tasks were independent samples t-tests. The schizophrenia group scored significantly lower on the digit symbol coding task compared to controls (*t*(34) = 3.278, *p* = 0.002), significantly lower on BVMT (*t*(34) = 2.407, *p* = 0.022) and the category fluency task (*t*(34) = 2.582, *p* = 0.014) compared to controls. There was no significant difference between groups on digit span forward (BF_10_ = 0.590) or digit span backwards (BF_10_ = 0.519).

**Table 2:**
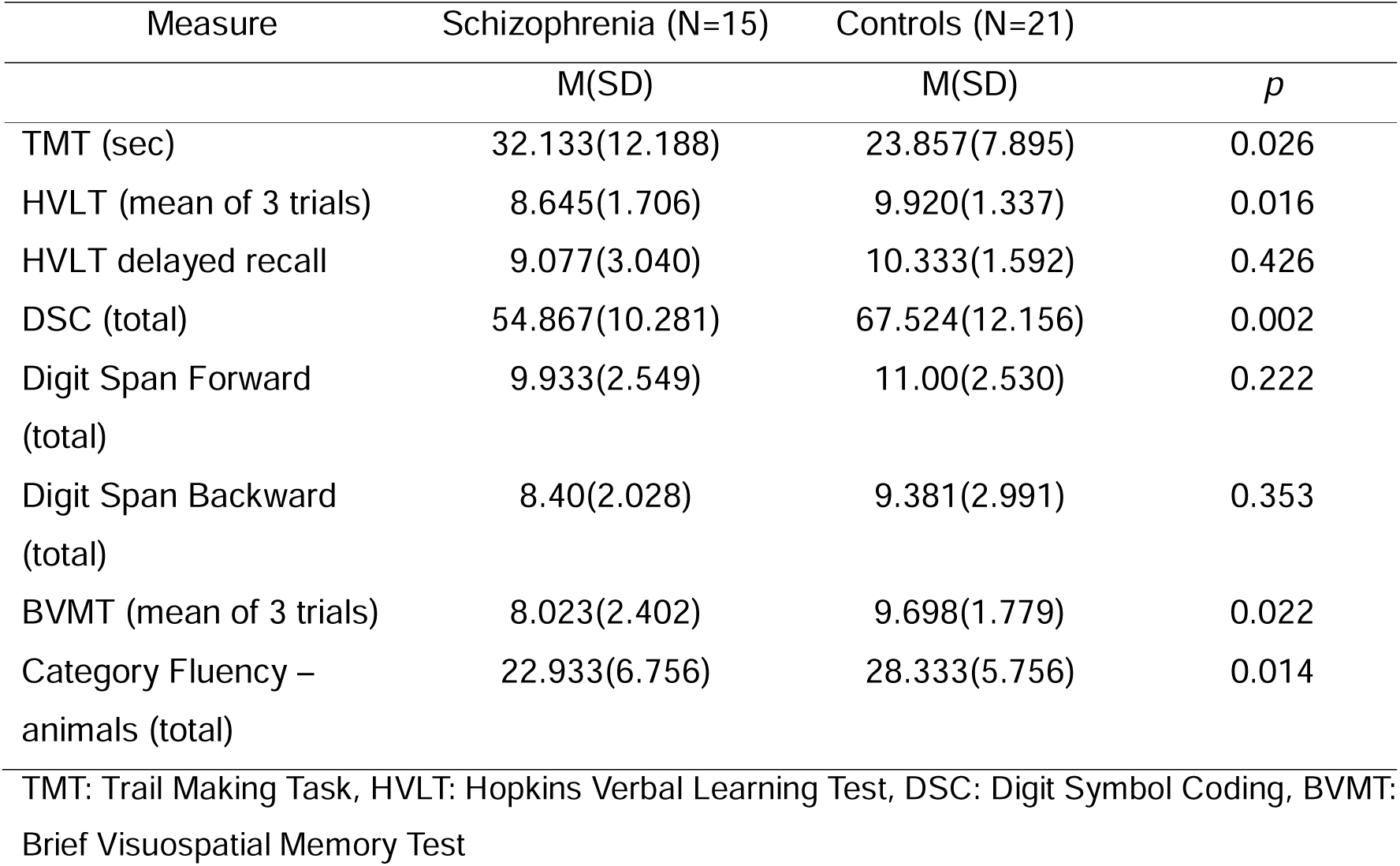
Cognitive Assessment Scores.

### 3.2 Theory of Mind Tasks

Participants with schizophrenia performed less well than controls on all ToM tasks across all assessment approaches. Specifically, on the traditional abbreviated faux pas recognition task, the schizophrenia group scored significantly lower compared to controls (*U* = 221.5, *p* = 0.035, r_rb_ = 0.41). On the computer-based attribution of intentions task, schizophrenia participants had significantly lower response accuracy on the ToM task (Welch, *t*(34) = 2.129, *p* = 0.045, *d* = 0.75) and NToM task (*t*(34) = 2.336, *p* = 0.026, *d* = 0.79) compared to the control group. The schizophrenia group also had a significantly slower response time on the ToM task (*t*(34) = −2.321, *p* = 0.026, *d* = −0.79) and NToM task (*t*(34) = −2.712, *p* = 0.010, *d* = −0.92) compared to controls. ERPs were used to further investigate the findings and are discussed in Section 3.4. Finally, on the VR ToM tasks, the schizophrenia group scored significantly lower on the RMET ToM task (Welch, *t*(18.978) = 2.508, *p* = 0.021, *d* = 0.89) and ToM hinting task (*U* = 260.50, *p* = <0.001, r_rb_ = 0.65). Z-scores were calculated to allow visual comparisons across the different tasks. Box-Cox transformations were used where violations of normality and/or variance were present (**Figure 3**). See supplementary materials for further details.

**Figure 3:**
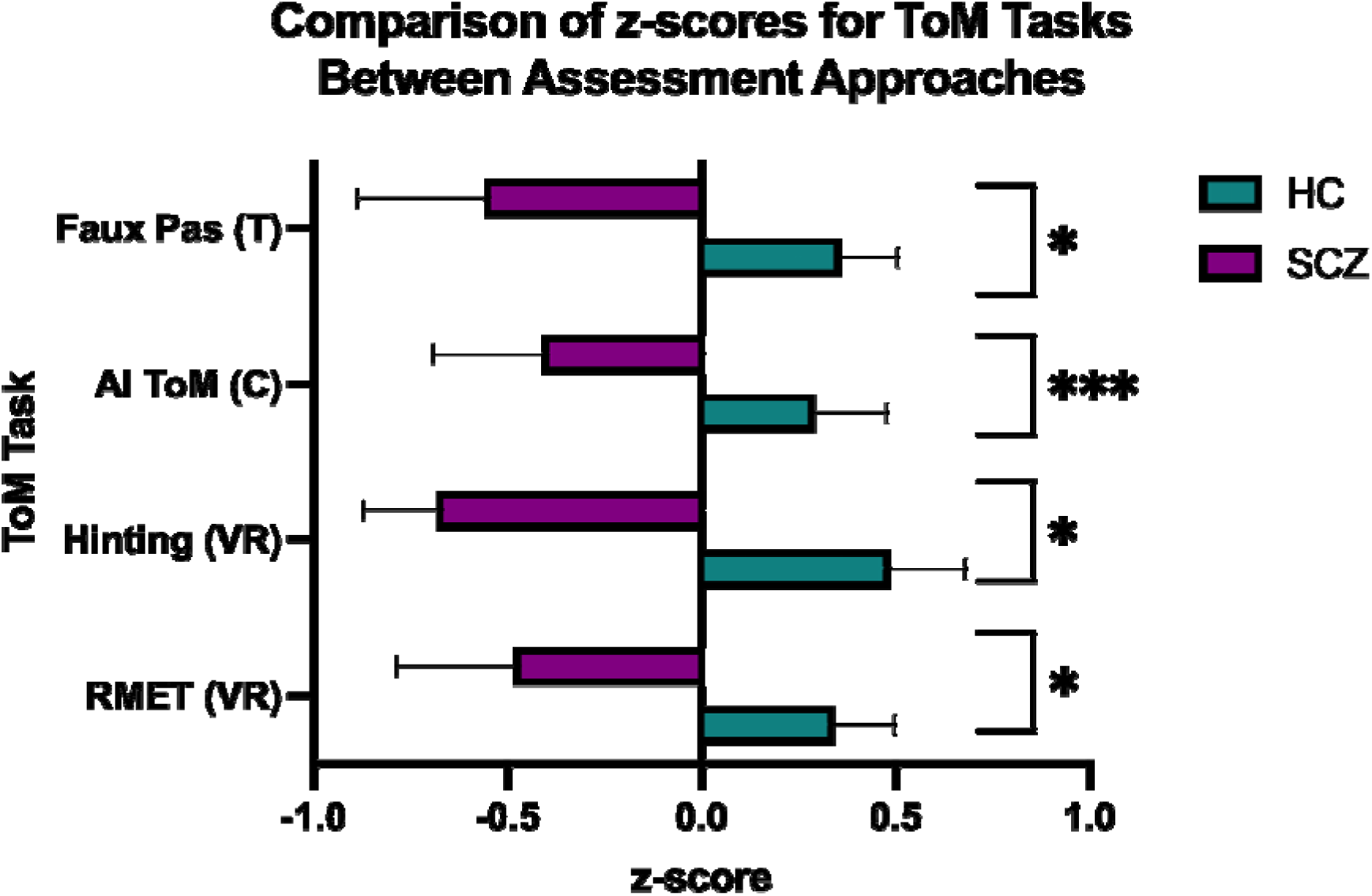
Comparison of z-scores between assessment approaches on ToM tasks. Note: C = computer task, VR = virtual reality task, T = traditional task. AI = attribution of intentions task, RMET = reading the mind in the eyes task. Significance levels: \**p* = < 0.05, \*\*\**p* = < 0.001.

### 3.3 Emotion Recognition, Social Knowledge and Attributional Bias

As with the ToM results, to allow direct comparison of results, z-scores were calculated to allow visual comparisons across the different tasks. Box-Cox transformations were used where violations of normality and/or variance were present (**Figure 4**). See supplementary materials for further details.

**Figure 4:**
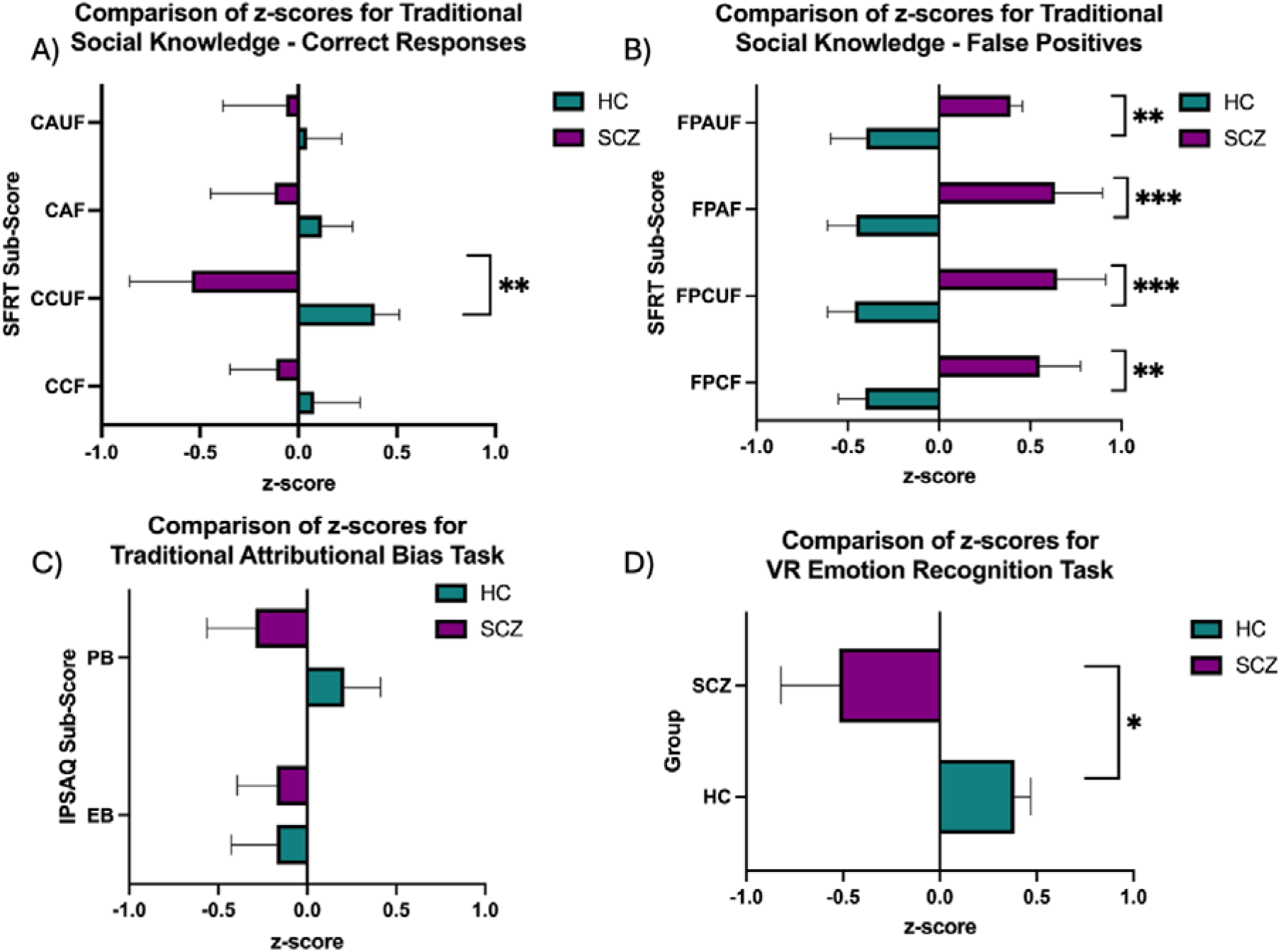
Comparison of task performance between schizophrenia and controls across assessment approaches using z-scores. Figure 4A) Represents sub-scores for correct responses on the situational features recognition test (SFRT), a traditional measure of social knowledge (CAUF = correct concrete features of unfamiliar situations, CAF = correct abstract features of familiar situations, CCUF = correct concrete features of unfamiliar situations, CCF = correct concrete features of familiar situations). Figure 4B) Shows representative scores for sub-scores of false positives on each of the aforementioned sub-scores (FPAUF = false positive abstract unfamiliar, FPAF = false positive abstract familiar, FPCUF = false positive concrete unfamiliar, FPCF = false positive concrete familiar. Figure 4C) Shows z-scores representative of the traditional IPSAQ sub-scores (PB = personalising bias, EB = externalising bias). Figure 4D) The customised emotion recognition task was conducted in VR. \**p* = <0.05, \*\**p* = <0.01, \*\*\**p* = <0.001.

#### 3.3.1 Emotion Recognition

The VR emotion recognition task showed a significant difference in mean scores between schizophrenia and controls with the schizophrenia group scoring significantly lower (*U* = 227.50, *p* = 0.025, r_rb_ = 0.44, **Figure 4D)**.

#### 3.3.2 Social Knowledge

On the traditional social knowledge measure (i.e., SFRT), significance scores varied. The schizophrenia group were less able to correctly identify concrete features of unfamiliar social situations (*U* = 236.50, *p* = 0.01, r_rb_ = 0.50, **Figure 4A**). They also had a higher false positive rate for identifying concrete features of familiar social situations (Welch, *t*(21.334) = −2.987, *p* = 0.007, *d* = −1.048), for identifying concrete features of unfamiliar social situations (*U* = 53.50, *p* = <0.001, r_rb_ = −0.66), for identifying abstract features of familiar situations (*U* = 51.00, *p* = <0.001, r_rb_ = −0.68) and for identifying abstract features of unfamiliar situations (*U* = 69.00, *p* = 0.004, r_rb_ = −0.56, **Figure 4B**). There was no significant difference between groups in the ability to correctly identify concrete features of familiar social situations (*U* = 185.00, *p* = 0.369, r_rb_ = 0.18, BF_10_ = 0.359), or identify abstract features of familiar social situations (*U* = 162.50, *p* = 0.882, r_rb_ = 0.03, BF_10_ = 0.646) as well as when identifying abstract features of unfamiliar social situations (*U* = 156.00, *p* = 0.974, r_rb_ = −0.01, BF_10_ = 0.395, **Figure 4A**).

#### 3.3.3 Attributional Bias

There was no significant difference between groups in the traditional attributional bias measure (IPSAQ) of externalising bias (*U* = 172.00, *p* = 0.65, r_rb_ = 0.09, BF_10_ = 0.524) or personalising bias (*t*(34) = 1.504, *p* = 0.142, *d* = 0.51, BF_10_ = 0.774, **Figure 5C**).

**Figure 5:**
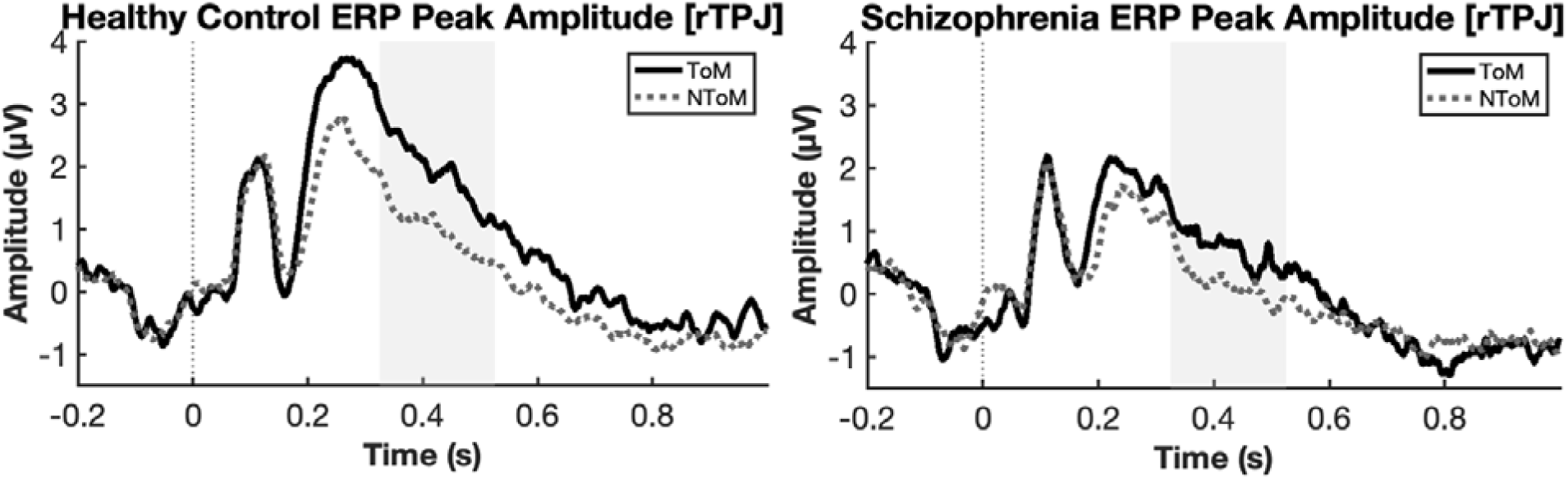
Comparison of ERP amplitudes during ToM and NToM tasks for schizophrenia sample and healthy controls. The grey vertical shading indicates the time point of interest for the TP450 ERP (i.e., 325ms-535ms).

### 3.4 Event-Related Potentials

A repeated measures ANOVA was conducted to investigate changes in TP450 ERP brain activity during ToM computer task performance (see Section 3.2) between schizophrenia and controls (IVs: group [healthy controls vs schizophrenia], task condition [ToM and NToM]; DV = mean ERP amplitude) at the ROI representing the right TPJ (CP4, CP6, TP8, P6, P8, **Figure 5**). There was a main effect of task condition (F(1, 34) = 17.839, *p* = <0.001, partial eta-squared = 0.344) with mean amplitude higher during ToM compared to NToM (mean difference = 0.745, SE = 0.176). There was also a main effect of group (F(1, 34) = 5.128, *p* = 0.030, partial eta-squared = 0.13) with mean amplitude higher for controls than for the schizophrenia group (mean difference = 1.019, SE = 0.450).

## 4. Discussion

This study investigated whether multiple assessment approaches could consistently detect a difference in social cognitive skills, specifically ToM, between people with a diagnosis of schizophrenia and healthy controls within the same sample. Findings supported the hypothesis by consistently demonstrating that in traditional, computer-based, and VR measures of ToM, participants with schizophrenia performed less well than healthy controls, with effect sizes for VR tasks being larger than traditional tasks. There was also reduced TP450 amplitude at the rTPJ for participants with schizophrenia compared to controls regardless of whether they were completing a ToM or NToM task and higher TP450 amplitude when completing ToM tasks compared to NToM regardless of group. There were also significant differences between groups on emotion recognition and social knowledge tasks.

### 4.1 Theory of Mind

Consistent with the literature that identifies ToM as one of the core social cognitive deficits in people with schizophrenia (Thibaudeau et al., 2021), the findings showed reduced ToM task performance compared to controls across all measures. However, the current study provides a novel approach that goes beyond usual assessment protocols by consistently demonstrating these ToM differences across various modalities within the same sample. Knowing these measures can sensitively detect differences between groups is useful for research and protocol development as it allows researchers to be confident that findings using any of these measures are robust effects. The current findings also suggest that the VR tasks may be more sensitive measures than traditional tasks. Indeed, results showed that the RMET VR task had the largest effect size (*d* = 0.89), followed by the ToM computer task (*d* = 0.75), VR hinting task (*d* = 0.65, moderate effect sizes) and lastly the traditional faux pas recognition task with a small effect size (r_rb_ = 0.41). As a result, when developing protocols, this hierarchy could guide outcome measurement selection during protocol development based on study requirements, access to equipment or participant abilities. For example, because the finding shows VR measures are the most sensitive, and therefore have more chance of accurately detecting effects, one might opt for this. However, if not available or the population being tested are less likely to be able to adequately operate the VR device, computer tasks or traditional methods could still be used with confidence given they were also able to sensitively detect a difference between groups.

The current findings are particularly encouraging for using VR to assess ToM skills. VR is increasingly being used in research and treatment for social cognition (Hoşgelen et al., 2024) because of its immersive qualities, ecological validity, and ability to tailor environments to needs (Parsons, 2015). It is possible that the ToM tasks being conducted in VR, were more able to tap into true ToM skills compared to the traditional task as they more closely represent real-world scenarios. Indeed, other studies adapting traditional cognitive tasks to virtual environments have shown similar results where participants found VR tasks more engaging and life-like compared to traditional versions (Johora, 2024; Zhou et al., 2023). While the current research uses tasks that are not entirely representative of social settings, (they display the standardised ToM tasks on a television in a comfortable living room setting), these findings are still promising and warrant further investigation using more socially engaging settings and directly comparing the same or similar ToM tasks in VR and traditional format.

### 4.2 Other Social Cognitive Domains

The current research also used other measures of cognition and social cognition to more broadly characterise our participant groups. We saw an expected deficit in cognitive performance as well as reduced performance in emotion recognition and social knowledge tasks. As with the ToM tasks, we used a novel VR measure of emotion recognition. Alongside the evidence for the ToM VR measures to detect a difference between groups, the finding supports VR as a good potential measure for other social cognitive domains. However, emotion recognition was assessed using only one approach and as such, further research using other emotion recognition tasks across assessment approaches may provide further robust, reliable evidence of the sensitivity of this measure.

Across all measures, the only assessment that failed to detect a difference between groups was the attributional bias measure (i.e., IPSAQ). While the IPSAQ is one of the most frequently used measures of attributional bias, a recent umbrella review (Samson et al., 2024) showed that the IPSAQ had a very small to no effect size when comparing schizophrenia and controls on both externalising and personalising bias measures even when sample sizes were considered good, no publication bias was detected, and confounding variables were absent (Samson et al., 2024). The measure is also self-report and while instructions are given, the two types of external attributions required (i.e., attribution to others or to the situation) can be misattributed and “water down” responses (Müller, Betz, & Bechdolf, 2021). This indicates that this finding may have potentially been influenced by the type of measure being used and could be further investigated by using an alternative measure of attributional bias in a similar multi-method protocol as the current study. Given the current findings showing VR-adapted measures had higher sensitivity to detecting effects, perhaps further investigation with VR measures could also be of value.

### 4.3 EEG as an Assistive Tool

The current paper also included an EEG measure of ERP (TP450) brain activity responses during ToM computer task performance to provide additional context to findings. There was a distinct neural activity pattern for the ToM-specific task in the rTPJ, a ToM hub (Masina et al., 2022; Wang, Callaghan, Gooding-Williams, McAllister, & Kessler, 2016), for both the schizophrenia and control groups. There was also an overall reduced activation in the rTPJ for people with schizophrenia regardless of whether they were completing the ToM or NToM task, suggesting an overall reduction of neural activity in ToM-relevant brain areas. However, although less activation, there is still a similar pattern of activity in the schizophrenia group as with healthy controls during the ToM task. This suggests that there is still specialised processing of ToM information but reduced activation for people with schizophrenia. These findings are in line with research by Vistoli, Brunet-Gouet, Lemoalle, Hardy-Baylé, and Passerieux (2011) using the same ToM task, showing brain activity in participants with schizophrenia followed the same temporal pattern with reduced activation at the rTPJ during the ToM task compared to controls. Taken together, these findings support the use of the rTPJ as a neural target for improving ToM performance. These findings are one example of how the use of neurophysiological measures such as EEG can be utilised alongside other social cognitive assessment approaches. Given the ToM findings of the current study, further research incorporating VR measures would be beneficial to identify neural biomarkers or treatment targets.

## 5. Limitations and Future Directions

While there was a significant difference between the schizophrenia and control groups in almost all assessments, only the ToM domain was tested across all assessment approaches. To allow even more robust interpretation of results, future studies would benefit from measuring all domains across all assessment approaches. The current study also used some novel assessments for measuring ToM which included the abbreviated Faux Pas from the Mini-SEA, less commonly used for schizophrenia samples (Karri, Bhavanani, Ramanathan, & Mopidevi, 2024), and the VR-adapted versions of the well-validated traditional RMET and Hinting task. The consistently significant ToM results suggest these measures are robust, however, future research with further use of these task versions in people with schizophrenia would further confirm this. The current study also selected tasks such as the IPSAQ that have, since being added to the protocol, been identified in other research as not being a sensitive enough measure (Samson et al., 2024). Conducting future research with multi-method protocols like the current one could again assist in validating these assessments further.

Additionally, the current findings suggest VR measures could be more sensitive than traditional measures; indicated by larger effect sizes for the former. However, these differences in effect sizes could also be a result of differences in ability to use VR technology for the participants with schizophrenia compared to paper and pencil tasks. Anecdotally, however, participants with schizophrenia were able to operate the VR with ease, as the tasks only required a single button press for each response. There was also no significant difference in groups regarding experience with the device when surveyed as part of the larger study. There was also a significant difference between the schizophrenia and control groups in age, the sample was not matched on sex between groups and overall, the sample size was small. While these differences are reflective of the general difficulty of recruiting participants in the community with a diagnosis of schizophrenia, these factors could have confounded the research and impacted results. Additionally, Bayes Factor scores were under one for non-significant findings. While this could potentially indicate adequate power, it does not rule out the possibility of low power in the study, especially in small samples such as this one (Stefan, Gronau, Schönbrodt, & Wagenmakers, 2019). Future research should ensure sample sizes are larger and are age and sex matched. Finally, combining the different assessment approaches used in the study in other ways could provide further, more robust and nuanced insights into symptom severity, potential treatment targets and efficacy of interventions. The current study administered EEG and computer-tasks at the same time but combining approaches in other ways such as VR and EEG, could allow even further advancement of sensitively measuring and identifying differences between groups as well as identifying treatment targets.

## 6. Conclusion

The current study used traditional, computer-based, and novel VR assessments to assess social cognition within the same sample. Across all assessment methods, a consistent reduction in social cognitive skills was observed in the schizophrenia group compared to controls. These findings provide support for using various types of measures to identify differences in social cognition between groups. Additionally, novel measures such as VR could be more sensitive than others and could be used on their own to reliably detect these differences or in combination with technologies such as EEG to further understand group differences. Introducing protocols like these could have implications for future treatment and protocol development by assisting with identifying treatment targets and accurately and robustly measuring symptom severity and changes for participants.

## Data Availability

All data produced in the present study are available upon reasonable request to the authors.

## Acknowledgements

We gratefully acknowledge the time and involvement from the participants. This research would not have been possible without them.

