## Supplementary Table 1 for "Comparing traditional, computerised and virtual reality assessments of social cognition in schizophrenia: A within-subjects multimodal approach"

1. Results

1.1 Box-Cox Transformations

**Table 1:** *Parameters used for Box-Cox data transformations in z-score comparisons*

| | Measure | Healthy Controls | Schizophrenia |
|---|---|---|---|
| | | Lambda(shift) | Lambda(shift) |
| ToM | | | |
| | Faux Pas | 7(0) | 2(0) |
| | Attribution of Intentions | 2.5(0) | 2.5(0) |
| | RMET | 2(0) | 2(0) |
| | Hinting Task | 4.5(0) | 4.5(0) |
| Emotion Recognition | | | |
| | VR Emotion Recognition | -3.5(0) | 3.5(0) |
| Social Knowledge (SFRT) | | | |
| | CCF | 2.5(0.001) | 2.5(0.001) |
| | CCUF | 3(0.001) | 3(0.001) |
| | CAF | 5(0.001) | 7(0.001) |
| | CAUF | 4(0.001) | 4(0.001) |
| | FPCF | No transform | 0.5(0.001) |
| | FPCUF | 0.5(0.001) | 0.5(0.001) |
| | FPAF | 0.3(0.001) | -0.3(0.001) |
| | FPAUF | | |
| Attributional Bias | | | |
| | Personalising Bias | No assumptions violated | No assumptions violated |
| | Externalising Bias | 0(3) | 2(8) |

Note: Data transformations were completed in JASP (Version 0.17.3, JASP Team, Amsterdam, Netherlands 2023) and parameters selected via visual inspection of distribution graphs. Code: ToM = Theory of Mind, RMET = Reading the Mind in the Eyes Task, SFRT = Situational Features Recognition Task, CCF = Correct Concrete Familiar, CCUF = Correct Concrete Unfamiliar, CAF = Correct Abstract Familiar, CAUF = Correct Abstract Unfamiliar, FPCF = False Positive Concrete Familiar, FPCUF = False Positive Concrete Unfamiliar, FPAF = False Positive Abstract Familiar, FPAUF = False Positive Abstract Unfamiliar.
